# Association between Glycemic Traits and Delayed Cerebral Infarction among Non-Diabetic Patients with Aneurysmal Subarachnoid Hemorrhage: A Nested Case-Control Study

**DOI:** 10.64898/2026.07.18.26358375

**Authors:** Peipei Ji, Kaijia Zheng, Dianhui Tan, Jincheng Xu, Muyuan Chen, Yanchun Wu, Zhen He

**Affiliations:** Department of Neurosurgery, First Affiliated Hospital of Shantou University Medical College, No.57 Changping Road, Shantou, Guangdong 515041, China; Shantou University Medical College, No.22 Xinling Road, Shantou, Guangdong 515041, China; Office of Nursing Research Institute, First Affiliated Hospital of Shantou University Medical College, No.57 Changping Road, Shantou, Guangdong 515041, China; Research Center of Translational Medicine, Department of Cardiology, Second Affiliated Hospital of Shantou University Medical College, No.69 Dongxia North Road, Shantou, Guangdong 515000, China

**Keywords:** aneurysmal subarachnoid hemorrhage, delayed cerebral infarction, glycemic variability, blood glucose

## Abstract

**OBJECTIVE:** Delayed cerebral infarction (DCIn) is a severe complication following aneurysmal subarachnoid hemorrhage (aSAH). Previous studies suggest that glycemic variability is associated with DCIn. However, whether diabetes status modifies the relationship between glycemic traits and DCIn remains unknown.

**METHODS:** Clinical data were collected from aSAH patients admitted to the First Affiliated Hospital of Shantou University Medical College between January 2015 and April 2025. The collected data included demographic characteristics, clinical variables, and glycemic traits. Glycemic traits included mean blood glucose (GLU-M), standard deviation of blood glucose (GLU-SD), coefficient of variation of blood glucose (GLU-CV), variance of blood glucose (GLU-Var), range of blood glucose (GLU-R), average real variability of blood glucose (GLU-ARV), and variability independent of the mean (GLU-VIM). After 1:2 case-control matching, conditional logistic regression models were used to evaluate the associations between glycemic traits and DCIn risk, with stratified analyses performed according to diabetes status. Multiplicative interaction terms were additionally included to assess the potential modifying effect of diabetes status.

**RESULTS:** A total of 306 patients with aSAH were included. Among them, 102 developed DCIn cases. For each of these 102 cases, two controls were matched by age (± 5 years), sex and year of admission (±5 years). In the overall population, higher GLU-M and GLU-ARV were associated with increased DCIn risk, with odds ratios (ORs) per 1-SD increase of 1.62 (95% CI, 1.25–2.11) and 1.63 (95% CI, 1.25–2.11), respectively. Among patients without diabetes (n=266), the associations with DCIn per 1-SD were observed for GLU-M (OR, 2.23; 95% CI, 1.56-3.19), GLU-SD (OR, 1.53; 95% CI, 1.13–2.06), GLU-Var (OR, 1.48; 95% CI, 1.04-2.10), and GLU-ARV (OR, 1.88; 95% CI, 1.38-2.55). No significant associations were observed among patients with diabetes. Significant interactions were observed between diabetes status and GLU-SD and GLU-Var, with *P* for interaction values of 0.033 and 0.032, respectively.

**CONCLUSION:** Higher mean blood glucose and greater glycemic variability are associated with an increased risk of DCIn in aSAH patients, especially in those without diabetes.

## Introduction

Aneurysmal subarachnoid hemorrhage (aSAH) is a severe subtype of hemorrhagic stroke, with a crude annual incidence of approximately 6.1 per 100,000 person-years^[1]^. Following aneurysm rupture, blood extravasation into the subarachnoid space leads to abrupt increases in intracranial pressure and reductions in cerebral perfusion, as well as triggers multiple pathological processes, including cerebral edema, blood-brain barrier disruption, impaired cerebral autoregulation, microthrombosis, spreading depolarization, and inflammation. As the disease progresses, subarachnoid blood and its degradation products may further aggravate these injury responses, ultimately leading to delayed cerebral ischemia (DCI)^[2–4]^. DCI develops in roughly 30% of aSAH patients within 4–14 days after rupture, and may progress to delayed cerebral infarction (DCIn)^[5,6]^, the most severe secondary complication, which results in severe disability or death in approximately 50% of affected patients^[7]^. However, the underlying mechanisms of DCIn remain unclear and warrant further investigation.

Glycemic variability (GV), defined as fluctuations in blood glucose levels, is typically quantified by measuring changes in glucose or related parameters over a specified time interval, including intraday, interday, or long-term periods^[8]^. It has become a more clinically relevant measure of glycemic control than HbA1c, and is being increasingly recognized for its importance^[9]^. The detrimental effects of elevated GV on target organs are mediated by oxidative stress, endothelial dysfunction, and chronic inflammation^[10]^. Emerging evidence underscores the harmful impact of pronounced glucose fluctuations, and a growing number of studies have consistently demonstrated strong associations between GV and adverse clinical outcomes, including increased risk of mortality^[11,12]^, cardiovascular events^[13–16]^, acute kidney injury^[17]^, cognitive impairment^[18]^, peripheral arterial disease^[19,20]^, osteoporotic fractures^[21]^ and digestive cancer^[22]^. Notably, the significance of glycemic variability may differ across different glycemic states^[11,14,19,21,23]^, suggesting that diabetes status should be considered when evaluating the association between GV and outcomes. Previous studies on GV in patients with aSAH have mainly focused on its predictive value for prognosis^[24–26]^, whereas studies specifically examining the relationship between GV and DCIn, and whether this relationship is modified by diabetes status, are relatively limited.

The aim of this study was therefore to investigate the effects of GV on DCIn among aSAH patients and to examine the role of diabetes status therein. To achieve this, we conducted a nested case-control study involving 306 aSAH patients with at least two blood glucose records, using data from the Department of Neurosurgery, the First Affiliated Hospital of Shantou University Medical College, between 2015 and 2025.

## Methods

### Study Population

Patient data were retrospectively collected from the Department of Neurosurgery in the First Affiliated Hospital of Shantou University Medical College between January 2015 and April 2025. Inclusion criteria were as follows: patients ≥18 years old; first-ever onset of symptoms, such as sudden severe headache or altered consciousness leading to emergency admission, subarachnoid hemorrhage confirmed by head CT and further verified as aneurysmal in origin by CT angiography (CTA) or digital subtraction angiography (DSA)^[27,28]^, and availability of at least two blood glucose measurements during hospitalization. Exclusion criteria were as follows: secondary subarachnoid hemorrhage (SAH) caused by trauma or other non-aneurysmal etiologies, a prior history of major neurological disorders that could influence outcome assessment, hospitalization lasting ≤ 24 hours, cerebral ischemic or infarction events (e.g., those caused by coil migration or escape, or parent artery stenosis after stent placement), and fewer than two blood glucose measurements obtained on different days, as well as patients with missing data. After applying these criteria, a final sample size of 306 patients was included (**Figure 1**). This study was conducted in accordance with the Declaration of Helsinki and approved by the Ethics Committee of the First Affiliated Hospital of Shantou University Medical College (approval number: B-2022-283). Informed consent was waived due to the retrospective nature of the study.

**Figure 1.**
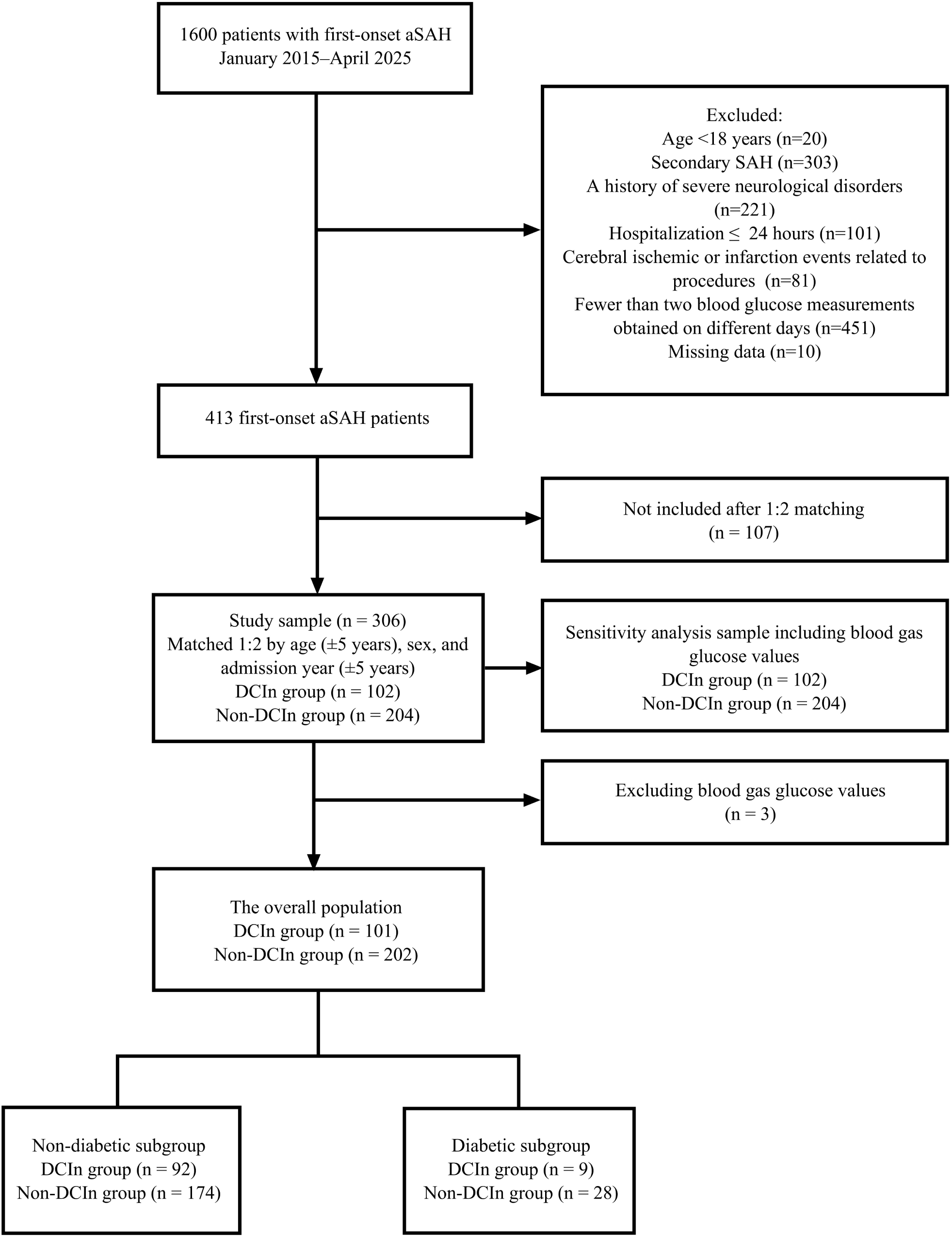
Flow chart of patient inclusion. aSAH, aneurysmal subarachnoid hemorrhage; DCIn, delayed cerebral infarction.

### Measurements of Traits

The trait variables, were glycemic traits, and included both mean blood glucose (GLU-M) and blood glucose variability. GLU-M was defined as the mean of all blood glucose measurements, while blood glucose variability was assessed using the following metrics: (1) standard deviation of blood glucose (GLU-SD)^[29]^, (2) coefficient of variation of blood glucose (GLU-CV), defined as GLU-SD divided by GLU-M^[29]^, (3) range of blood glucose (GLU-R), calculated as the difference between the maximum and minimum blood glucose values, (4) variance of blood glucose (GLU-Var), defined as the mean of the squared deviations of glucose measurements from their mean, (5) average real variability of blood glucose (GLU-ARV), defined as the average absolute difference between adjacent blood glucose measurements^[13]^, (6) variability independent of the mean of blood glucose (GLU-VIM), calculated as 100 × SD/mean^^β^, where β is the regression coefficient obtained from the regression of the natural logarithm of GLU-SD over the natural logarithm of the mean^[30]^. The formulas for these indices are provided below.

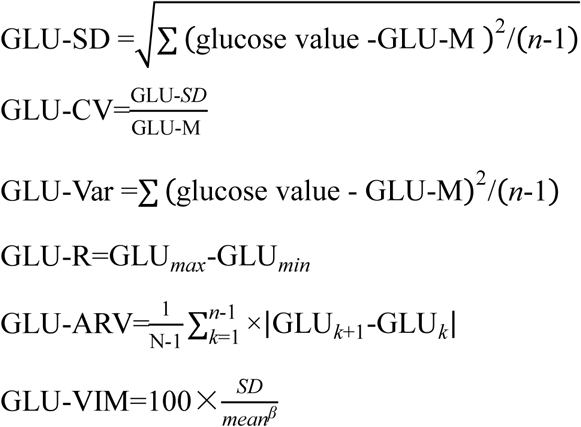

Blood glucose values included both random and fasting measurements, as well as glucose values obtained from blood gas analysis using arterial blood. In Table 1, we present the descriptive data of participants who met the inclusion criteria, including 102 cases with DCIn, and 204 age-matched controls. However, in the multivariable models (e.g., Tables 2–5), we excluded participants whose glucose levels were measured using arterial blood, as we believe such measurements are not comparable to those obtained from venous blood. Nevertheless, we included glucose values measured from arterial blood in the sensitivity analyses (Tables S9–S14). For aSAH patients who developed DCIn during hospitalization, only blood glucose measurements obtained before DCIn onset were included, whereas for those who did not develop DCIn, all in-hospital blood glucose measurements were included.

**Table 1.**
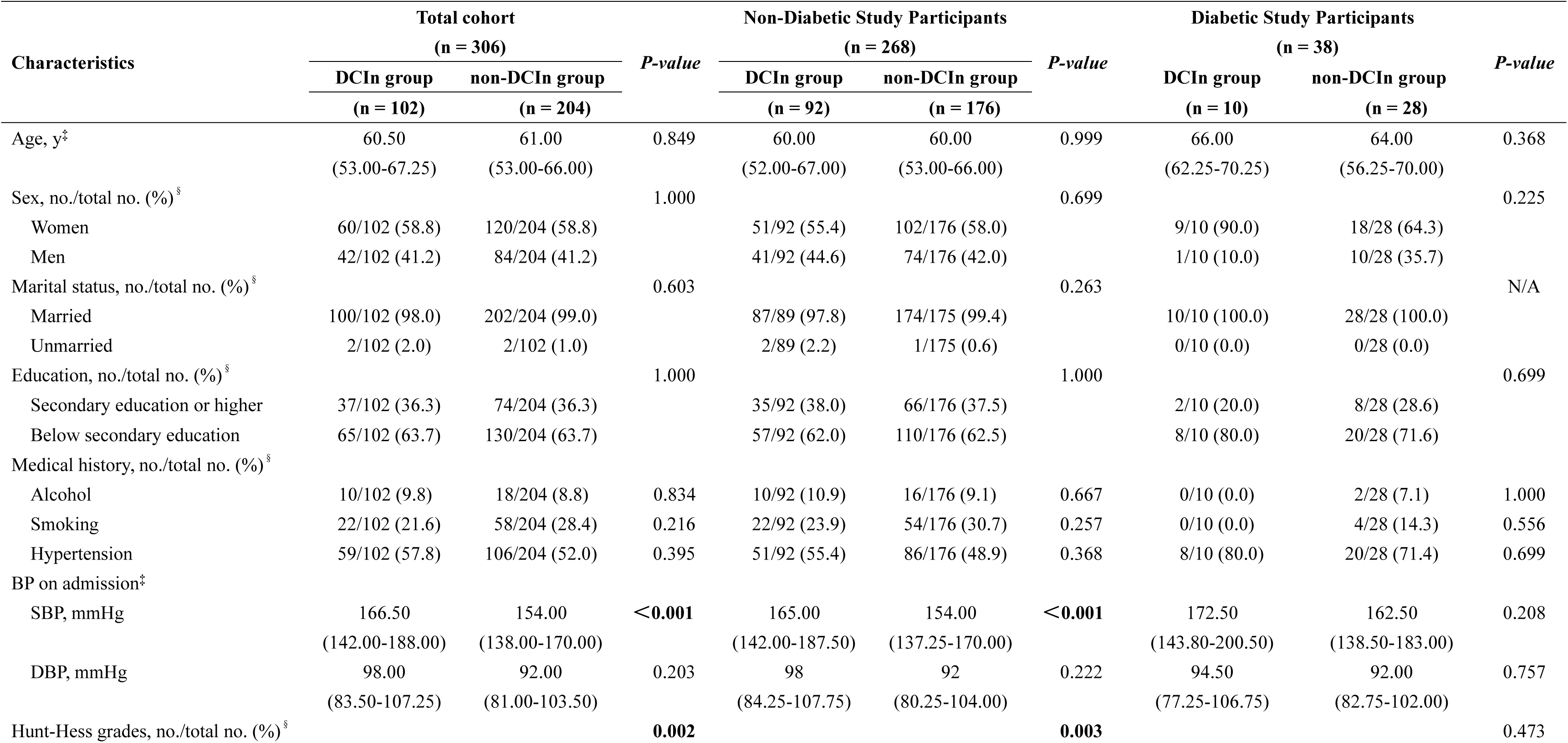
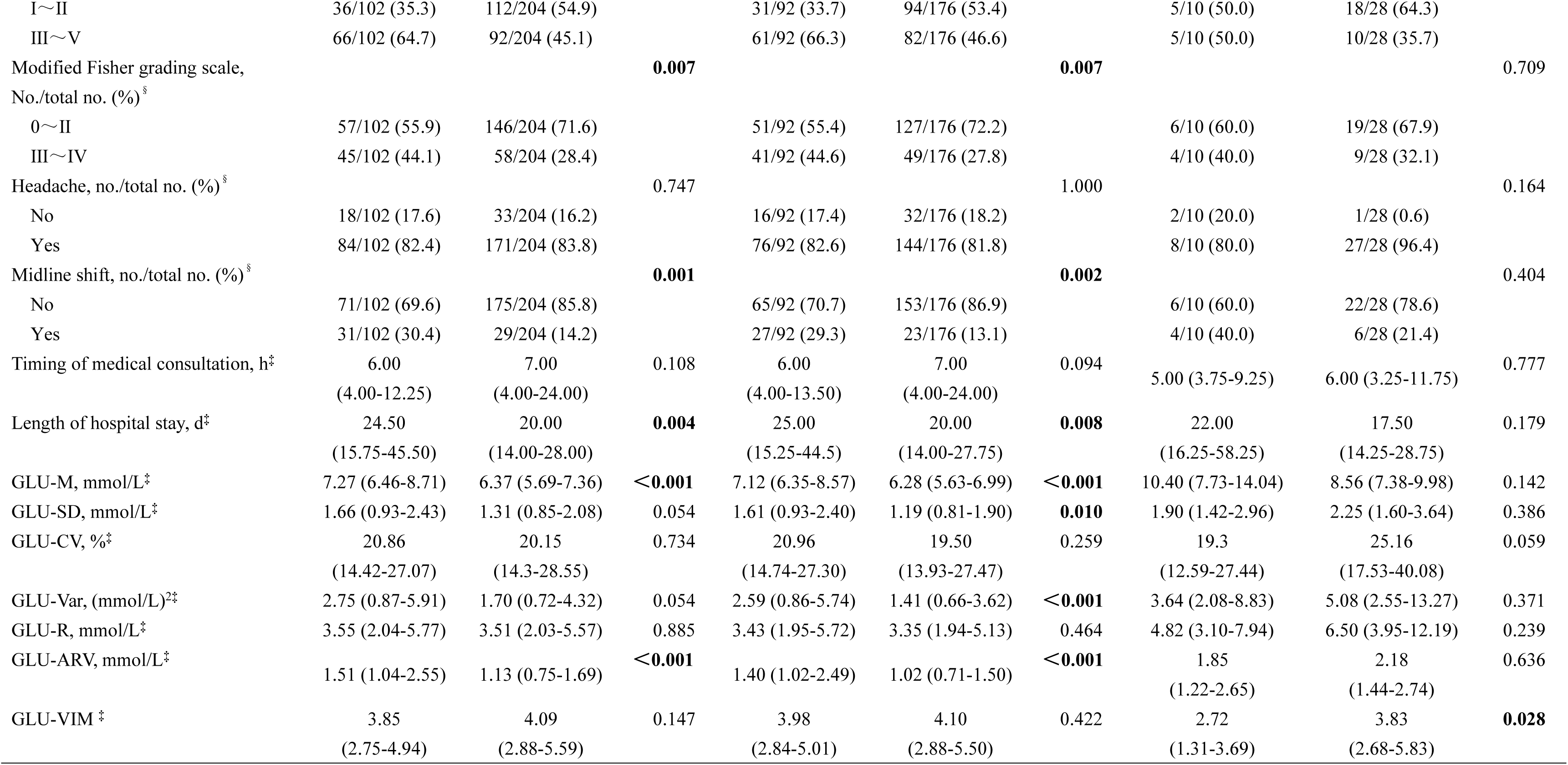
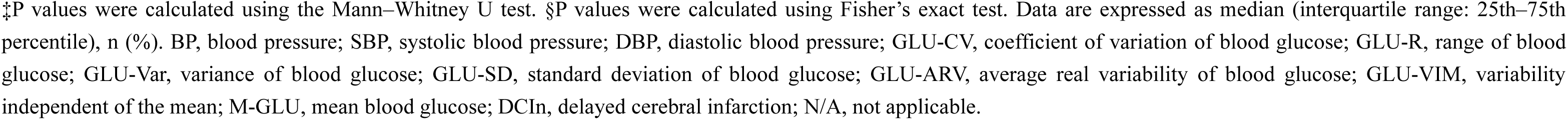
Baseline Characteristics of Participants Stratified by Diabetes Status and DCIn Categories.

**Table 2.** Multivariable-adjusted associations of glycemic variability and GLU-M with DCIn risk in the overall population using conditional logistic regression without blood gas variables.

| Variable<br>(per 1 SD higher) | DCIn group (n=101) vs Non-DCIn group (n=202) |  |  |  |  |  |  |  |
| --- | --- | --- | --- | --- | --- | --- | --- | --- |
|  | OR (95% CI) | *P-value* | OR (95% CI) | *P-value* | OR (95% CI) | *P-value* | OR (95% CI) | *P-value* |
|  | Model1 |  | Model2 |  | Model3 |  | Model4 |  |
| GLU-M | 1.60 (1.24-2.07) | **<0.001** | 1.63 (1.26-2.11) | **<0.001** | 1.62 (1.25-2.11) | **<0.001** | 1.49 (1.14-1.96) | **0.004** |
| GLU-SD | 1.23 (0.97-1.56) | 0.094 | 1.26 (0.99-1.61) | 0.066 | 1.24 (0.97-1.60) | 0.087 | 1.18 (0.91-1.53) | 0.205 |
| GLU-CV | 1.06 (0.83-1.35) | 0.630 | 1.08 (0.85-1.38) | 0.514 | 1.07 (0.84-1.38) | 0.569 | 1.05 (0.81-1.36) | 0.713 |
| GLU-Var | 1.12 (0.88-1.41) | 0.363 | 1.13 (0.89-1.44) | 0.308 | 1.12 (0.87-1.43) | 0.380 | 1.07 (0.83-1.39) | 0.585 |
| GLU-R | 0.99 (0.77-1.28) | 0.958 | 1.02 (0.79-1.31) | 0.905 | 1.00 (0.77-1.28) | 0.970 | 0.89 (0.68-1.16) | 0.385 |
| GLU-ARV | 1.63 (1.25-2.12) | **<0.001** | 1.64 (1.26-2.13) | **<0.001** | 1.63 (1.25-2.11) | **<0.001** | 1.56 (1.20-2.02) | **0.001** |
| GLU-VIM | 0.87 (0.68-1.11) | 0.264 | 0.88 (0.69-1.13) | 0.322 | 0.88 (0.68-1.13) | 0.306 | 0.89 (0.68-1.16) | 0.375 |

Model 1: unadjusted;
Model 2: adjusted for age and sex;
Model 3: Model 2 plus educational level, alcohol consumption, smoking, and year of hospital admission;
Model 4: Model 3 plus history of hypertension, Hunt–Hess grade, and modified Fisher grading scale.
Abbreviations: OR, odds ratio. GLU-M, mean blood glucose; GLU-SD, standard deviation of blood glucose;
GLU-CV, coefficient of variation of blood glucose; GLU-Var, variance of blood glucose; GLU-R, range of blood
glucose; GLU-ARV, average real variability of blood glucose; GLU-VIM, variability independent of the mean;
DCIn, delayed cerebral infarction.

**Table 3.**
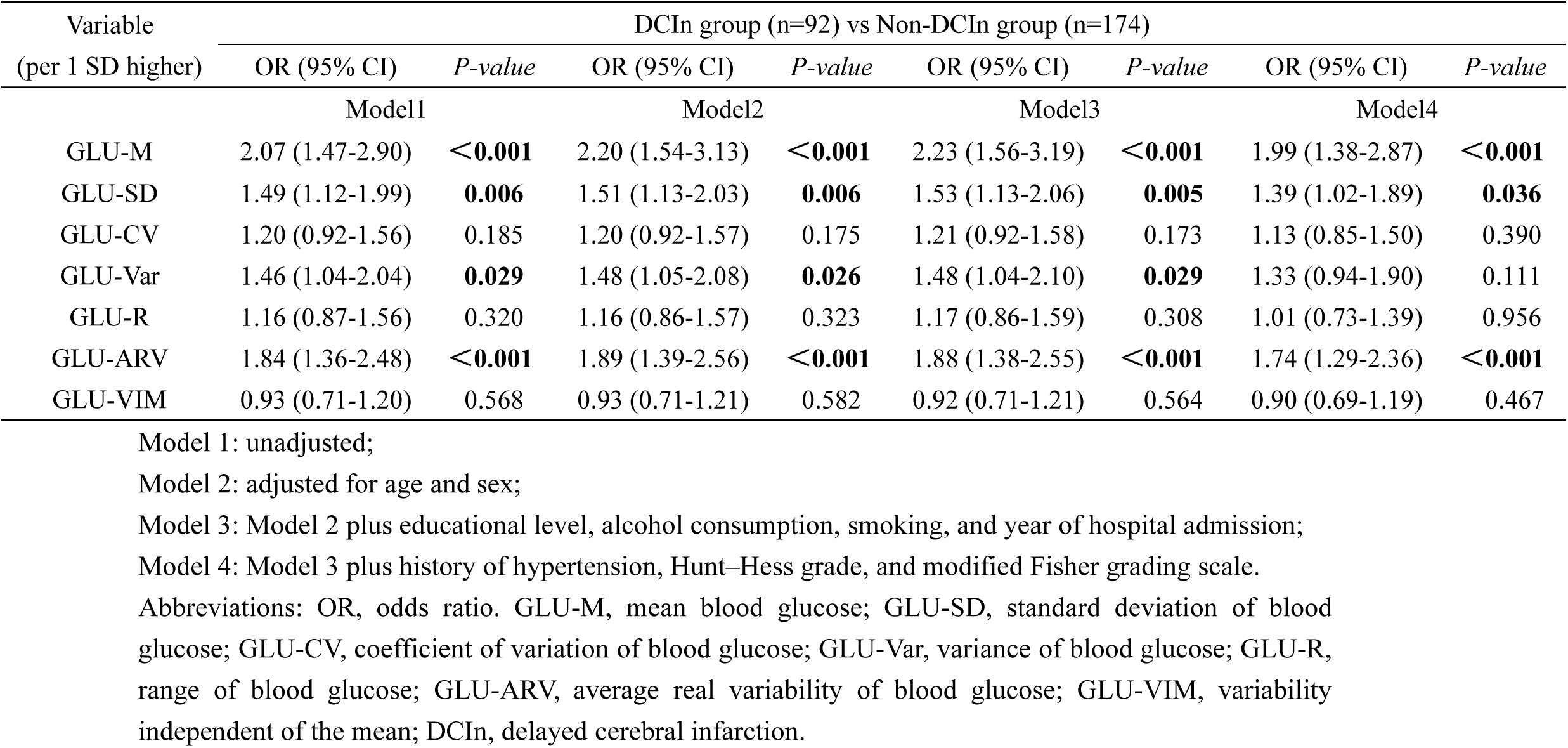
Multivariable-adjusted associations of glycemic variability and GLU-M with DCIn risk in non-diabetic patients using unconditional logistic regression without blood gas variables.

**Table 4.**
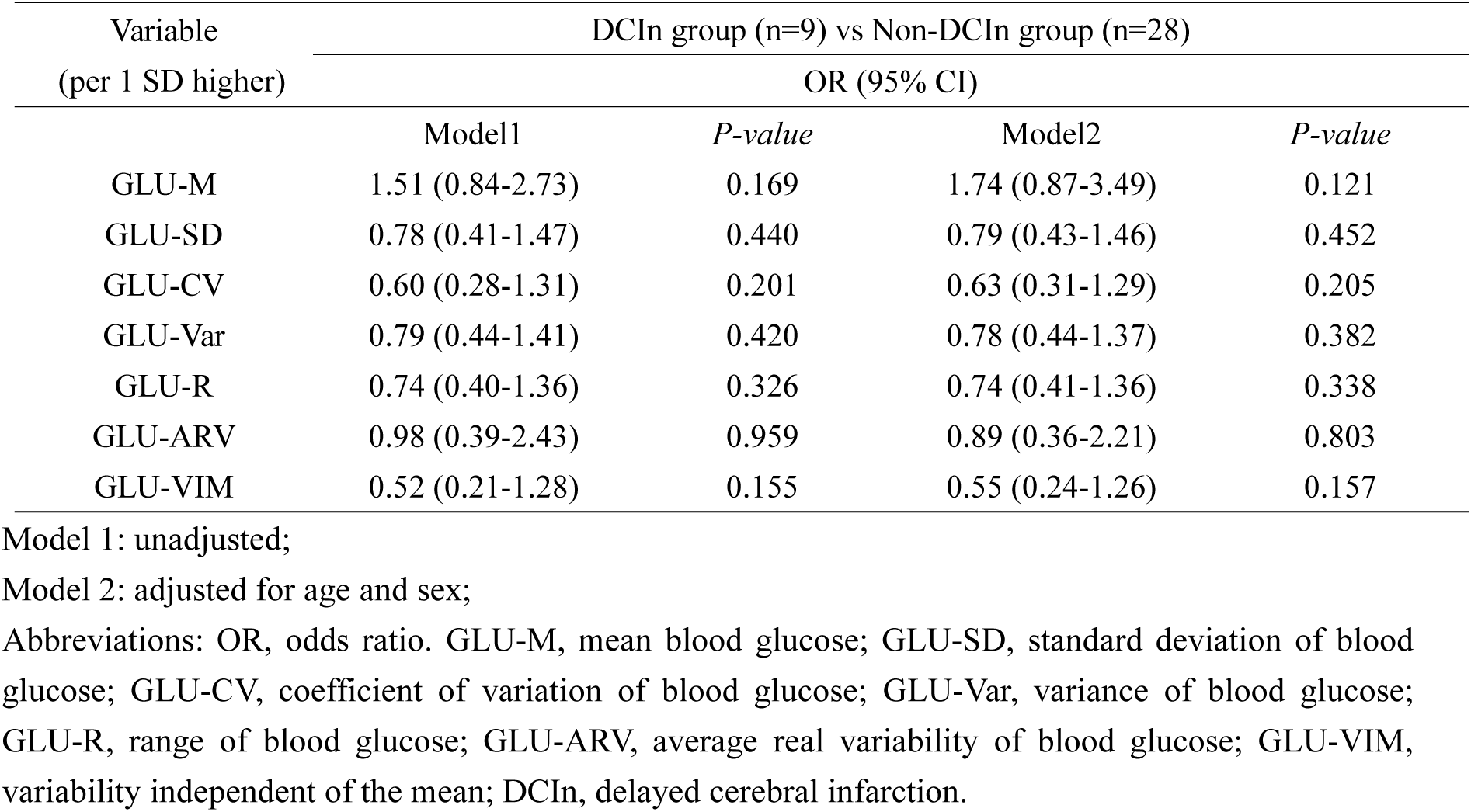
Multivariable-adjusted associations of glycemic variability and GLU-M with DCIn risk in diabetic patients using unconditional logistic regression without blood gas variables.

**Table 5.** Multivariable-adjusted interactions between glycemic variability, GLU-M, and diabetes status in relation to DCIn risk: conditional logistic regression analysis without blood gas variables.

| Variable | DCIn group (n=101) vs Non-DCIn group (n=202) |  |  |  |
| --- | --- | --- | --- | --- |
|  | OR (95% CI) | P-value | OR (95% CI) | P-value |
|  | Model 3 |  | Model 4 |  |
| GLU-M* | 2.17 (1.51-3.14) | <0.001 | 1.97 (1.37-2.85) | <0.001 |
| Diabetes | 0.33 (0.10-1.06) | 0.063 | 0.40 (0.12-1.34) | 0.139 |
| **GLU-M:Diabetes** | **0.63 (0.31-1.29)** | **0.208** | **0.62 (0.30-1.27)** | **0.194** |
| GLU-SD* | 1.57 (1.17-2.12) | 0.003 | 1.46 (1.08-1.99) | 0.015 |
| Diabetes | 0.63 (0.24-1.63) | 0.340 | 0.71 (0.27-1.89) | 0.493 |
| **GLU-SD:Diabetes** | **0.46 (0.22-0.94)** | **0.033** | **0.47 (0.23-0.97)** | **0.042** |
| GLU-CV* | 1.25 (0.95-1.64) | 0.119 | 1.19 (0.89-1.58) | 0.239 |
| Diabetes | 0.65 (0.27-1.58) | 0.344 | 0.68 (0.27-1.71) | 0.410 |
| **GLU-CV:Diabetes** | **0.43 (0.18-1.02)** | **0.056** | **0.46 (0.19-1.14)** | **0.093** |
| GLU-Var* | 1.55 (1.09-2.21) | 0.015 | 1.44 (1.01-2.06) | 0.044 |
| Diabetes | 0.62 (0.25-1.52) | 0.296 | 0.70 (0.27-1.79) | 0.456 |
| **GLU-Var:Diabetes** | **0.47 (0.23-0.94)** | **0.032** | **0.48 (0.23-0.97)** | **0.042** |
| GLU-R* | 1.23 (0.90-1.67) | 0.200 | 1.06 (0.76-1.48) | 0.724 |
| Diabetes | 0.68 (0.27-1.69) | 0.402 | 0.80 (0.31-2.08) | 0.645 |
| **GLU-R:Diabetes** | **0.56 (0.29-1.11)** | **0.095** | **0.59 (0.30-1.19)** | **0.139** |
| GLU-ARV* | 1.86 (1.38-2.51) | <0.001 | 1.73 (1.28-2.33) | <0.001 |
| Diabetes | 0.52 (0.19-1.38) | 0.186 | 0.57 (0.21-1.56) | 0.273 |
| **GLU-ARV:Diabetes** | **0.49 (0.18-1.37)** | **0.176** | **0.54 (0.18-1.61)** | **0.271** |
| GLU-VIM* | 0.95 (0.72-1.25) | 0.707 | 0.94 (0.71-1.24) | 0.638 |
| Diabetes | 0.50 (0.19-1.29) | 0.150 | 0.51 (0.19-1.37) | 0.184 |
| **GLU-VIM:Diabetes** | **0.47 (0.17-1.31)** | **0.149** | **0.54 (0.18-1.60)** | **0.268** |

Model 3: adjusted for age, sex, educational level, alcohol consumption, smoking, and year of hospital admission;
Model 4: Model 3 plus history of hypertension, Hunt–Hess grade, and modified Fisher grading scale.
\*, per 1 SD higher; OR, odds ratio; GLU-M, mean blood glucose; GLU-SD, standard deviation of blood glucose; GLU-CV, coefficient of variation of blood glucose; GLU-Var, variance of blood glucose; GLU-R, range of blood glucose; GLU-ARV, average real variability of blood glucose; GLU-VIM, variability independent of the mean; DCIn, delayed cerebral infarction.

Venous blood glucose levels were measured using an Abbott glucose assay kit based on the hexokinase/glucose-6-phosphate dehydrogenase (HK/G6PD) method. Venous blood samples were obtained by standard venipuncture, collected into coagulation tubes, and centrifuged to obtain serum for glucose determination. Arterial blood gas glucose was measured using a fully automated blood gas analyzer (GEM Premier 3500, Werfen, Spain), which determines whole-blood glucose concentration, using the electrochemical electrode method.

### DCIn Diagnosis

Delayed cerebral infarction was defined as the presence of cerebral infarction on CT or MR imaging, appearing within 6 weeks after aSAH or on the last CT or MR scan obtained before death within 6 weeks, that was not present on the CT or MR scan performed between 24 and 48 hours after early aneurysm occlusion, and that could not be attributed to other causes, such as surgical clipping or endovascular treatment. Hypodensities on CT resulting from ventricular catheter placement or intraparenchymal hematoma were not considered cerebral infarctions related to DCI^[31]^.

### Covariates

According to the epidemiological definition of confounding factors, the included variables were age, sex, year of hospital admission (continuous variable), alcohol (current/non-current), smoking (current/non-current), education level (below secondary vocational education/secondary vocational education or above), history of hypertension (yes/no), Hunt-Hess grades (I – II/III – V), and modified Fisher grading scales (0 – II/III – IV).

Hypertension was defined as self-reported hypertension, the use of antihypertensive medications, or a systolic/diastolic blood pressure ≥ 140/90 mmHg^[32]^. Diabetes was defined as self-reported physician-diagnosed diabetes, the use of antidiabetic medication, or a fasting blood glucose level ≥ 7.0 mmol/L^[33]^. The Hunt– Hess grading scale was used to assess clinical aSAH severity at admission, and ranged from grade I to grade V, with higher grades indicating more severe neurological impairment. Specifically, grades I – II were classified as low-grade aSAH, and grades III – V were classified as high-grade aSAH^[34]^. The modified Fisher grading scale was applied to assess the volume and distribution of intracranial hemorrhage on cranial CT and MRI, with grades 0 – II classified as the low-risk group, and grades III – IV as the high-risk group^[35]^. Midline shift was assessed on the admission CT scan and defined as the horizontal displacement of the septum pellucidum from the bony midline, which was determined by the line connecting the crista galli and the inion at the level of the foramen of Monro^[36]^. Smoking status was defined as having smoked at least 100 cigarettes in one’s lifetime and is currently smoking cigarettes at least on some days. Participants who did not meet both criteria were classified as non-current smokers^[37]^. Alcohol consumption was defined as having consumed at least one standard drink, equivalent to 10 g of pure ethanol, during the past 12 months. Participants who did not meet this criterion were classified as non-current drinkers^[38]^. Educational level, year of hospital admission and the presence of headache at onset were obtained from electronic medical records. Headache at onset was defined as a sudden thunderclap headache, characterized by an abrupt and unprecedented severe headache. Timing of commencement of medical consultation was defined as the interval between the time of symptom onset and arrival at our hospital.

### Statistical Analysis

Case-control matching was performed at a 1:2 ratio between the DCIn and non-DCIn groups, using age (± 5 years), sex, and year of hospital admission (± 5 years) as matching variables. We additionally considered matching by year of hospital admission because medical conditions, such as equipment, technology, reagents, and other factors, may vary over time.

Baseline characteristics were presented as median (interquartile range [IQR], 25th–75th percentile) for non-normally distributed continuous variables and number (percentage) for categorical variables. Continuous variables were compared using the Mann-Whitney U test, and categorical variables were compared between groups using Fisher’s exact test. Differences with a two-sided *P* < 0.05 were considered statistically significant.

Conditional logistic regression models were used to evaluate the associations between glycemic traits and the risk of DCIn. Analyses were conducted separately among participants with and without diabetes. Before analysis, all trait variables were standardized (mean = 0, SD = 1) to improve comparability. For each trait variable, including GLU-M, GLU-SD, GLU-CV, GLU-Var, GLU-R, GLU-ARV, and GLU-VIM, four models were constructed: Model 1 was unadjusted, Model 2 was adjusted for age and sex, Model 3 was additionally adjusted for educational level, alcohol consumption, smoking status and year of hospital admission, and Model 4 was further adjusted for history of hypertension, Hunt–Hess grade, and the modified Fisher grading scale. We considered Model 3 as the main analysis, as the variables adjusted in this model were likely potential confounders of the glycemic trait-DCIn relationship. Additionally, we added an interaction term (glycemic traits × diabetes status) to Models 3 and 4 to evaluate the modifying role of diabetes status.

To examine the robustness of the findings, several sensitivity analyses were conducted: (1) repeating the analyses using unconditional logistic regression models, (2) repeating the analyses using the original, unstandardized trait variables, (3) truncating excess glucose measurements among individuals with more frequent testing within each matched set to match the individual with fewer measurements, followed by repetition of the analyses, (4) including glucose values obtained from blood gas analysis and repeating the analyses, and (5) including blood gas-derived glucose values and additionally truncating excess measurements as described above, followed by repetition of the analyses.

The risk of DCIn was expressed as odds ratios (ORs) with 95% confidence intervals (CIs). All statistical analyses were performed using IBM SPSS Statistics software (version 27.0, IBM Corp., Armonk, NY, USA) and R (version 4.5.2, R Foundation for Statistical Computing, Vienna, Austria). Conditional logistic regression analyses were performed using the survival package in R, whereas unconditional logistic regression analyses were performed using the glm function in R. This study was conducted in accordance with the STROBE guidelines for case-control studies (Supplementary File 1. STROBE Statement Checklist)^[39]^.

## Results

### Baseline Characteristics

Three hundred six patients with aSAH were included. Among them, 102 patients (33.3%) developed DCIn. The median age was 61 years, and 58.8% were women. The median interval from blood glucose variability assessment to the onset of DCIn was 25.3 hours. Compared with the non-DCIn group, patients in the DCIn group had higher systolic blood pressure (median [IQR], 166.50 [142.00 – 188.00] vs. 154.00 [138.00 – 170.00] mmHg), higher Hunt-Hess (66/102 [64.7%] vs. 92/204 [45.1%]), and modified Fisher grades (45/102 [44.1%] vs. 58/204 [28.4%]), greater midline shift (31/102 [30.4%] vs. 29/204 [14.2%]), longer hospital stays (median [IQR], 24.50 [15.75–45.50] vs. 20.00 [14.00–28.00] days), and higher GLU-M (median [IQR], 7.27 [6.46 – 8.71] vs. 6.37 [5.69 – 7.36] mmol/L), and GLU-ARV levels (median [IQR], 1.51 [1.04–2.55] vs. 1.13 [0.75–1.69] mmol/L) (all *P* < 0.05). No significant differences were observed in age, sex, marital status, educational level, alcohol consumption, smoking status, history of hypertension, diastolic blood pressure, headache, timing of medical consultation, or other glucose variability indices between the two groups (all *P* > 0.05) (Table 1). Similarly, the non-diabetic subgroup showed patterns largely consistent with those of the overall population, except that GLU-SD and GLU-Var were higher in the DCIn group (both *P* < 0.05). However, GLU-VIM was the only parameter that showed a positive and significant association with DCIn in diabetic patients (Table 1).

### Associations of Glycemic Traits with DCIn Risk in the Overall Population

Among the overall population, after excluding glucose values from arterial blood, higher GLU-M was associated with a higher risk of DCIn in Model 3 (OR per 1 SD increase GLU-M, 1.62 [95% CI, 1.25-2.11]). A similar pattern of association was observed for GLU-ARV (OR per 1 SD increase, 1.63 [95% CI, 1.25-2.11]). In contrast, no statistically significant associations were observed for other glycemic variability indicators, including GLU-SD, GLU-CV, GLU-Var, GLU-R, and GLU-VIM (all *P* > 0.05; Model 3, Table 2 and Figure 2).

**Figure 2.**
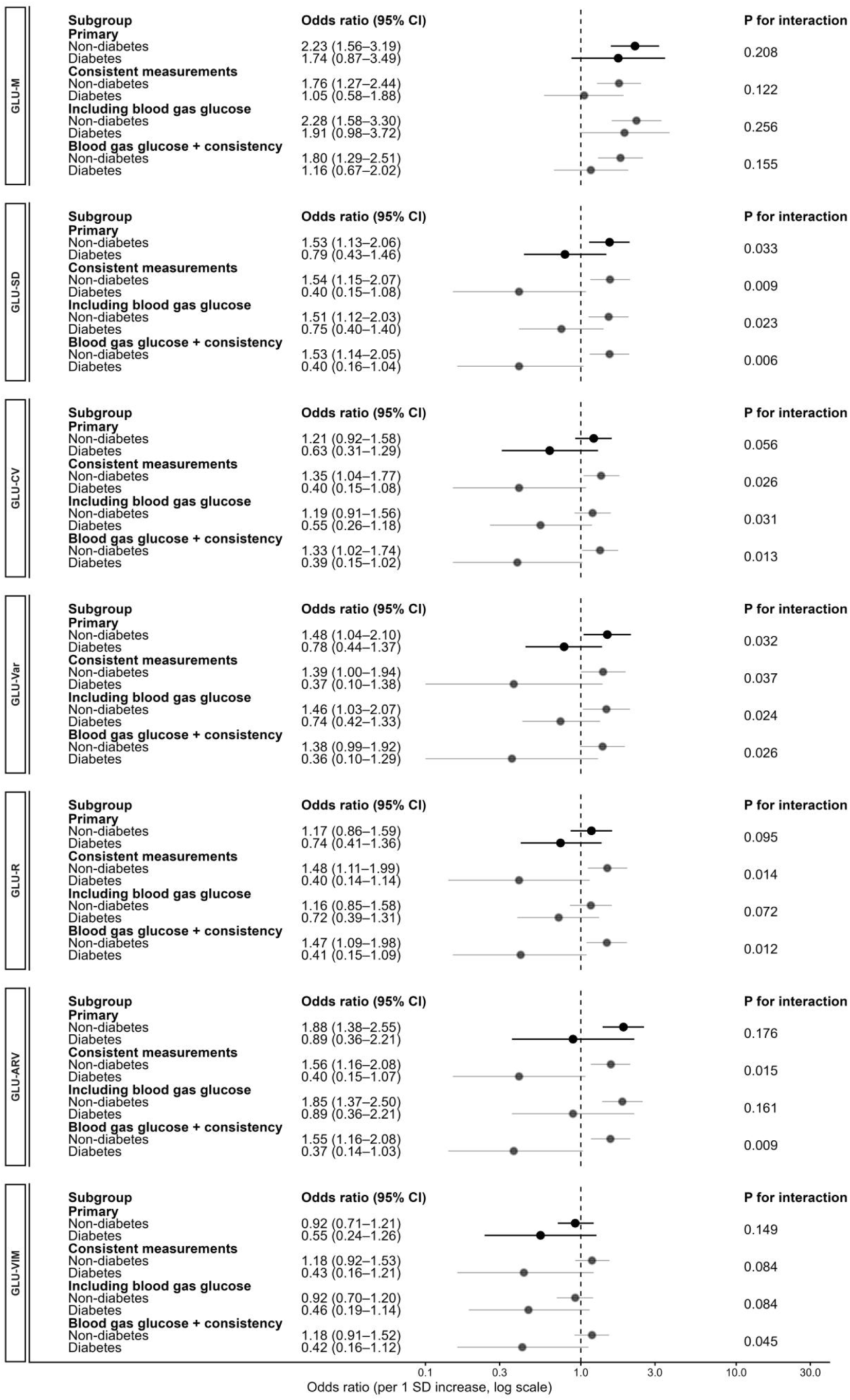
Association between glycemic indices and delayed cerebral infarction (DCIn) stratified by diabetes status. Forest plots show adjusted odds ratios (ORs) and 95% confidence intervals (CIs) for DCIn per 1-SD increase in different glycemic indices, stratified by diabetes status (non-diabetes vs diabetes). Dots indicate point estimates of ORs, and horizontal lines indicate 95% CIs. Black dots and corresponding horizontal lines indicate the primary analysis, whereas gray dots and corresponding horizontal lines indicate sensitivity analyses. The vertical dashed line indicates an OR of 1.0. Panels correspond to individual glycemic measures, including mean glucose level (GLU-M), standard deviation of glucose (GLU-SD), coefficient of variation of glucose (GLU-CV), variance of glucose (GLU-Var), range of glucose (GLU-R), average real variability of glucose (GLU-ARV), and variability independent of the mean (GLU-VIM). Results are presented for the primary analysis excluding glucose values derived from blood gas analysis, and for sensitivity analyses, including analyses with a consistent number of glucose measurements within each matched set, analyses including blood gas–derived glucose values, and analyses including blood gas–derived glucose values with a consistent number of glucose measurements.

### Associations of Glycemic Traits with DCIn Risk in Non-diabetic Patients

In non-diabetic patients, by excluding glucose values from arterial blood, the overall trend was similar to that observed in the overall population. Both higher GLU-M (per 1 SD increase, 2.23 [95% CI, 1.56-3.19]) and GLU-ARV (per 1 SD increase, 1.88 [95% CI, 1.38-2.55]) were positively associated with DCIn. Additionally, GLU-SD (per 1 SD increase, 1.53 [95% CI, 1.13-2.06]) and GLU-Var (per 1 SD increase, 1.48 [95% CI, 1.04-2.10]) were also higher in the DCIn group, whereas GLU-CV, GLU-R, and GLU-VIM showed no significant differences between the two groups (all *P* > 0.05; Table 3). These findings were also highly consistent across Models 1, 2 and 4 (Table 3).

### Associations of Glycemic Traits with DCIn Risk in Diabetic Patients

Among participants with diabetes, after adjustment for age and sex, neither glycemic variability indices nor GLU-M levels were associated with the risk of DCIn (Model 2, Table 4, and Figure 2).

### Interactions between Glycemic Traits and Diabetes Status

In the interaction analysis, the terms for GLU-SD × diabetes status and GLU-Var × diabetes status were both statistically significant (*P* for interaction values were 0.033 and 0.032, respectively; Table 5, Figure 2). However, no significant associations were observed for the other interaction terms.

### Sensitivity Analyses

The primary findings shown in Tables 2-4 were corroborated by several sensitivity analyses, including models that balanced glucose measurement frequency (Table S6-S7), models using arterial blood data (Table S9-S10,S12-S13), and models based on unconditional logistic regression (Table S1). The interaction terms for GLU-SD × diabetes status and GLU-Var × diabetes status remained significant under sensitivity analyses that balanced measurement frequency and used arterial blood glucose values (Tables S8, S11, S14).

## Discussion

This study was a nested case–control study and included glycemic indices obtained prior to the occurrence of DCIn. The results show that among adult patients with aSAH, higher average GLU-ARV and higher GLU-M are associated with an increased risk of DCIn. However, diabetes status exerted a significant effect-modifying role on the association between glycemic parameters and the risk of DCIn. Specifically, higher M-GLU and greater glycemic variability were associated with an increased risk of DCIn only in patients without diabetes, whereas no significant association was observed in patients with diabetes. Sensitivity analyses further supported the robustness of the above findings.

Our findings are supported by several animal and observational studies. Huang et al. demonstrated in a rat SAH model that hyperglycemia exacerbates cerebral vasospasm, as evidenced by a markedly greater reduction in basilar artery cross-sectional area in the hyperglycemic SAH group compared with the SAH-only group^[40]^. Consistent with these experimental findings, Deininger et al., in a retrospective, single-center observational study of 151 aSAH patients, found that the time-weighted average glucose was already higher in patients who subsequently developed DCI than in non-DCI patients (128.7 vs. 124.3 mg/dL, p=0.024), with a further abrupt rise at the time of DCI onset, suggesting a temporal precedence of glycemic disturbance relative to ischemic events^[41]^. A previous retrospective study of severe SAH patients (n=42) further showed that a glucose change rate >9.52 mg/dL/h during the first 7 days is associated with a markedly higher incidence of cerebral infarction (64% vs. 20%), and remains an independent predictor after multivariable adjustment (non-standardized OR=11.4, 95% CI: 1.9-70.2), although its conclusions are limited by small sample size and inability to confirm the temporal relationship between glucose fluctuations and infarction^[42]^. However, this unusually large effect estimate should be interpreted cautiously, given the small sample size, wide confidence interval, potential overfitting related to the data-driven threshold and multivariable model, possible selection bias from including only patients with Hunt-Hess grade ≥3, and the inability to confirm the temporal relationship between glucose fluctuations and infarction. Qiu et al. subsequently applied group-based trajectory modeling (GBTM) to 2,182 aSAH patients, identifying four distinct glucose trajectory patterns over the first 14 days, with the persistently elevated and highly fluctuating trajectory associated with higher DCI risk compared with the low-stable trajectory (aOR=1.63, 95% CI: 1.09–2.43)^[43]^. The present study complements these findings by separately quantifying glycemic variability and mean glucose as independent variables, demonstrating that both are associated with higher DCIn risk, particularly in non-diabetic patients.

Several mechanisms may account for the observed association between glucose parameters and risk of DCIn in nondiabetics. From a physiological standpoint, diabetic patients may develop adaptive responses—often referred to as “metabolic preconditioning” — as a result of chronic exposure to glycemic fluctuations^[44,45]^. These adaptations may enhance their tolerance to acute glucose variability during aSAH. In contrast, nondiabetic patients, who typically maintain tighter glycemic control under normal conditions, lack such compensatory mechanisms. Consequently, their tissues, particularly endothelial cells, are more vulnerable to acute glycemic excursions, which can induce oxidative stress, exacerbate neuronal injury^[46]^, cause endothelial dysfunction that disrupts the integrity of the blood-brain barrier, and impair cerebral hemodynamic stability^[41,47]^. Moreover, glucose variability promotes the release of neuroinflammatory mediators and enhances immune cell infiltration, thereby amplifying the neuroinflammatory response^[48]^. Moreover, impaired regulation of cerebral energy metabolism secondary to glucose instability leads to an imbalance between energy supply and demand in neural tissue^[26]^. Collectively, these interrelated pathophysiological processes act synergistically to exacerbate secondary brain injury and contribute to the development of DCIn.

This study may still be subject to several potential sources of bias. First, the inclusion of only patients with available glucose monitoring data and at least two measurements, which may have introduced selection bias. In addition, the frequency and timing of glucose measurements were not fully consistent, and glucose values may differ according to blood sample source, which could have affected the estimation of glycemic variability. To mitigate these concerns, we performed sensitivity analyses by further balancing the number of glucose measurements between cases and controls, and by additionally incorporating arterial blood glucose values, and showed the results were generally consistent with the primary analysis. Second, the relatively long enrollment period may have introduced time-related bias due to changes in clinical techniques, laboratory equipment, reagents, and perioperative management strategies that occur over time. To address this issue, year of hospital admission was considered during the matching process to minimize the potential influence of temporal changes in medical treatments across different periods. Third, although this study used a matched case–control design, matching itself may not fully control for confounding introduced by the matching factors, as highlighted by Pearce et al^[49]^. Therefore, age, sex, and year of admission were still used for adjustment in the matched analyses. Unconditional logistic regression analyses were also performed to evaluate the robustness of the findings, which yielded consistent results. Fourth, we constructed four sequentially adjusted models to evaluate the robustness of associations between glycemic indicators and DCIn risk under varying levels of confounding control, demonstrating highly consistent results. Fifth, because we examined seven traits, we applied a Bonferroni correction for multiple testing (α = 0.00714 [0.05/7]). After correction, the associations of GLU-M and GLU-ARV with DCIn remained significant (both *P* < 0.001). Finally, no significant associations were observed in the diabetic subgroup, which may be attributable to insufficient statistical power due to the relatively small sample size. Accordingly, false negative findings cannot be excluded in this subgroup.

This study has several strengths. First, given the limited evidence on the relationship between glycemic traits and DCIn, this study provides a relatively systematic evaluation of the associations between multiple glycemic traits and the risk of DCIn in patients with aSAH. Second, the nested case–control design and the inclusion of only glycemic indices obtained before the occurrence of DCIn helped to clarify the temporal sequence between trait and outcome. In addition, the nested case–control design improved study efficiency and reduced resource consumption, while cases and controls were derived from the same cohort, thereby enhancing comparability between groups. Finally, sensitivity analyses were conducted, and the results were generally consistent with the primary analysis, supporting the robustness of our findings.

This study also has several limitations. First, this was a retrospective study, and some potential covariates (such as lifestyle habits and dietary information) could not be fully obtained; therefore, residual confounding cannot be completely excluded, and the ability to infer causality remains limited. Second, as an observational study design, the nested case–control study has generally lower statistical power than prospective cohort studies. Third, since glucose data were derived from routine clinical monitoring, differences in measurement frequency, timing, and blood sample source were inevitable, and measurement bias cannot be completely avoided. Fourth, this study was based on single-center data, and the findings may not be generalizable to other regions or different ethnic populations. Fifth, diabetes was defined based on acute-phase glucose levels rather than glycated hemoglobin in this study. Consequently, some non-diabetic patients may have been misclassified as having diabetes. Nevertheless, no association between glycemic traits and DCIn was observed among diabetic patients. The impact of this misclassification bias is difficult to quantify and would require larger cohorts of diabetic patients for further investigation. Finally, the relatively small sample size may have resulted in insufficient statistical power, particularly in the diabetic subgroup analysis.

In non-diabetic patients with aSAH, we observed that glycemic variability was associated with risk of DCIn, suggesting that acute-phase dynamic glucose changes may reflect disruption of metabolic homeostasis under stress, and may participate in the occurrence and progression of secondary ischemic brain injury. These findings highlight the need for more refined monitoring of glucose dynamics in this population. Compared with intermittent glucose monitoring, continuous glucose monitoring (CGM) can more comprehensively capture glycemic fluctuation patterns ^[50]^. Although CGM has been widely used in diabetes management, emerging evidence suggests its feasibility and favorable monitoring performance during the early brain injury phase in non-diabetic patients with SAH, enabling more sensitive assessment of early glycemic dysregulation^[51]^. Future prospective studies are warranted to evaluate whether CGM can facilitate early identification of patients at high risk of DCIn and guide individualized glucose management strategies, thereby optimizing perioperative glycemic management.

## Conclusion

In conclusion, higher GLU-M and greater glycemic variability are associated with an increased risk of DCIn among aSAH patients, particularly in those without diabetes. Future studies are warranted to elucidate the mechanisms underlying the interrelationships between glycemic variability measures and non-diabetic status in relation to the risk of DCIn in patients with aSAH, and to evaluate whether targeting these factors is an effective strategy for prevention of DCIn development. Moreover, larger studies are needed to further investigate the association between glycemic variability and DCIn, especially in diabetic populations.

## Conceptualization

Peipei Ji, Yanchun Wu, and Zhen He. Methodology: Peipei Ji, Kaijia Zheng, and Zhen He. Data curation: Peipei Ji and Kaijia Zheng. Formal analysis: Peipei Ji. Investigation: Peipei Ji, Kaijia Zheng, Dianhui Tan, Jincheng Xu, and Muyuan Chen. Writing-original draft: Peipei Ji. Writing-review and editing: Yanchun Wu, and Zhen He. Supervision: Yanchun Wu and Zhen He. All authors read and approved the final manuscript.

## Supporting information

Supplementary File 1. STROBE Statement Checklist

## Data Availability

The data that support the findings of this study are available from the corresponding author upon reasonable request.

## Acknowledgments

We would like to thank Stanely Lin for their valuable assistance with language editing.

## Notes

### Competing Interest Statement

The authors have declared no competing interest.

### Author Declarations

The Ethics Committee of the First Affiliated Hospital of Shantou University Medical College gave ethical approval for this work (approval No. B-2022-283).

