## Supplementary File 1. STROBE Statement Checklist for "Association between Glycemic Traits and Delayed Cerebral Infarction among Non-Diabetic Patients with Aneurysmal Subarachnoid Hemorrhage: A Nested Case-Control Study"

| | Item Number | Recommendation | Location in manuscript |
|---|---|---|---|
| Title and Abstract | 1 | (a)Indicate the study's design with a commonly used term in the title or the abstract. | Page 1, Title |
| | | (b)Provide an informative and balanced summary of what was done and what was found. | Page 1-2, Abstract |
| Introduction | | | |
| Background/rationale | 2 | Explain the scientific background and rationale for the investigation being reported. | Page 3-4, Background |
| Objectives | 3 | State specific objectives, including any prespecified hypotheses. | Page 4, Objectives |
| Methods | | | |
| Study design | 4 | Present key elements of study design early in the paper. | Page 4, Study design |
| Setting | 5 | Describe the setting, locations, and relevant dates, including periods of recruitment, exposure, follow-up, and data collection. | Page 4, Setting |
| Participants | 6 | (a)Cohort study: eligibility criteria, and sources/methods of participant selection; describe follow-up.<br>Case–control: eligibility criteria, and sources/methods of case ascertainment and control selection.<br>Cross-sectional: eligibility criteria, and sources/methods of participant selection.<br>(b)Cohort study: for matched studies, give matching criteria and number of exposed/unexposed.<br>Case–control: for matched studies, give matching criteria and number of controls per case. | Page 4, Participants |
| Variables | 7 | Clearly define outcomes, exposures, predictors, potential confounders, and effect modifiers. | Page 5-7, Variables |
| Data sources/ | 8[a] | For each variable, give data | Page 6-7, Data sources/ |

| | | | |
|---|---|---|---|
| measurement | | sources and methods of assessment;<br>describe comparability between groups. | measurement |
| Bias | 9 | Describe efforts to address potential sources of bias. | Page 8, Bias |
| Study size | 10 | Explain how the study size was arrived at. | |
| Quantitative variables Statistical methods | 11 | Explain how quantitative variables were handled; describe groupings if applicable. | Page 7-8, Quantitative variables |
| | 12 | (a) Describe all statistical methods, including confounding control.<br>(b) (b) Describe methods for subgroups and interactions.<br>(c) (c) Explain how missing data were handled.<br>(d) (d) Cohort: explain how loss to follow-up was addressed.<br>Case–control: explain how matching was addressed.<br>Cross-sectional: describe analytical methods accounting for sampling strategy.<br>(e) Describe any sensitivity analyses. | Page 8-9, Statistical methods |
| Results | | | |
| Participants | 13[a] | (a) Report numbers of individuals at each study stage (eligible, included, followed- up, analyzed). | Page 5, Results, Participants; Figure 1 |
| | | (b) Give reasons for non-participation. | Page 5, Results, Participants; Figure 1 |
| | | (c) Consider a flow diagram. | Page 5, Results, Participants; Figure 1 |
| Descriptive data | 14[a] | (a) Give participant characteristics (demographic, clinical, social) and information on exposures/confounders.<br>(b) Indicate number of participants with missing data for each variable.<br>(c) Cohort study: summarise follow-up time (e.g., average and total). | Page10, Descriptive data |

| | | | |
|---|---|---|---|
| Outcome data | 15 | Cohort: report outcome events or summary measures over time.<br>Case–control: report numbers in each exposure category.<br>Cross-sectional: report outcome events or summary measures. | Page10-11, Outcome data |
| Main results | 16 | (a) Give unadjusted and, if applicable, adjusted estimates with precision (e.g., 95% CI); state confounders adjusted for.<br>(b) Report category boundaries when continuous variables were categorized.<br>(c) Translate estimates of relative risk into absolute risk when relevant. | Page10-11, Main results |
| Other analyses | 17 | Report other analyses (subgroups, interactions, sensitivity analyses). | Page10-11, Report other analyses |
| Discussion | | | |
| Key results | 18 | Summarize key results with reference to study objectives. | Page11, Key results |
| Limitations | 19 | Discuss study limitations and potential bias (direction and magnitude). | Page14, Limitations |
| Interpretation | 20 | Interpret results cautiously considering objectives, limitations, multiplicity of analyses, and other evidence. | Page11-14, Interpretation |
| Generalisability | 21 | Discuss generalizability of study findings. | Page14-15, Generalisability |
| Other information | | | |
| Funding | 22 | Give source of funding and the role of funders. | |

[a] Give such information separately for cases and controls in caseecontrol studies, and, if applicable, for exposed and unexposed groups in cohort and cross-sectional studies. Note: An Explanation and Elaboration article discusses each checklist item and gives methodological background and published examples of transparent reporting. The STROBE checklist is best used in conjunction with this article (freely available on the Web sites of PLoS Medicine at http://www.plosmedicine.org/, Annals of Internal Medicine at http://www.annals.org/, and Epidemiology at http://www.epidem.com/). Separate versions of the checklist for cohort, caseecontrol, and cross-sectional studies are available on the STROBE Web site at http://www.strobe-statement.org/.
